# Therapygenetics: Serotonin transporter gene polymorphism (*5-HTTLPR*) is associated with behavioral treatment response to virtual exposure therapy in patients with spider phobia

**DOI:** 10.1101/2025.06.26.25330346

**Authors:** Elisabeth Schrammen, Chiara Hilbrich, Joscha Böhnlein, Kati Roesmann, Bettina Gathmann, Martin J. Herrmann, Markus Junghöfer, Hanna Schwarzmeier, Fabian R. Seeger, Niklas Siminski, Thomas Straube, Heike Weber, Ulrike Lueken, Udo Dannlowski, Katharina Domschke, Miriam A. Schiele, Elisabeth J. Leehr

## Abstract

Identifying biomarkers predicting therapy outcomes before treatment holds great promise for advancing precision medicine. Genetic variants such as the serotonin transporter gene linked polymorphic region (*5-HTT*LPR) may be associated with response to cognitive behavioral therapy in anxiety disorders, albeit results so far are controversial. Contributing to the ongoing debate, we investigated whether treatment response to a highly standardized one-session virtual reality exposure therapy (VRET) was predicted by *5-HTT*LPR genotype. N=194 patients with arachnophobia (spider phobia) were genotyped for *5-HTT*LPR and the functionally related single nucleotide polymorphism rs25531 and grouped into high-(L_A_/L_A_), and low-expression (S/S, S/L_G,_ L_G_/L_G_, S/L_A,_ L_G_/L_A_) genotype. At baseline, after VRET, and at a 6-month follow-up, participants underwent a standardized behavioral avoidance task (BAT) and the spider phobia questionnaire (SPQ) to assess symptom severity. Chi-square tests revealed a significant association between *5-HTT*LPR/rs25531 and behavioral treatment outcome, that remained significant at the 6-month follow-up. No association was found between genotype and self-reported symptom severity measured with the SPQ. Our results support the idea that while L_A_/L_A_genotype carriers might benefit from highly standardized treatment, lower *5-HTT* expression may convey risk to poorer treatment response, likely necessitating more tailored psychotherapeutic interventions to promote sufficient response.

## 1 Introduction

Anxiety disorders have the highest prevalence among mental disorders and convey a significant individual and socioeconomic burden (Wittchen et al., 2011). Within the group of anxiety disorders, specific phobias are the most common (Craske et al., 2017). Despite their prevalence, specific phobias are rarely accompanied by comorbid disorders or treated with psychotropic drugs, rendering them an ideal model with strong internal validity for studying the mechanisms underlying the pathogenesis and treatment response in anxiety disorders. The recommended first-line treatment for specific phobias is exposure therapy. Even short interventions – most studies report one session of one to three hours length – have been shown to be effective (e.g., Leehr et al., 2021). Exposure therapy can either be delivered *in vivo* or via virtual reality (VRET), with comparable treatment outcome (Wechsler et al., 2019). Despite its documented high efficacy, roughly two thirds of treated patients do not respond to exposure treatment, failing to achieve a clinically significant outcome (Cuijpers et al., 2024). Recently, there has been a growing interest in the identification of biomarkers of (psycho-)therapeutic success. By identifying the right patients for the right treatments, healthcare providers can offer more precise, effective, and safe medical care, ultimately improving patient outcomes and the overall efficiency of the healthcare system (Lueken & Hahn, 2016). Numerous candidate risk genes have been put forward. Among those, one of the most studied genes is the serotonin receptor gene (*5-HTT*), and, more precisely, the serotonin transporter gene-linked polymorphic region (*5-HTT*LPR), a length polymorphism located within the gene’s promotor region (Ansorge et al., 2004; Wendland et al., 2006). *5-HTT*LPR comprises a high-expressing long (L) and a low-expressing short (S) allele (Hariri & Holmes, 2006; Lesch et al., 1996). A single nucleotide polymorphism (rs25531, A>G) within the L-allele further influences 5-HTT expression, with presence of the rs25531 G allele rendering it functionally equivalent to the S-allele (Wendland et al., 2006). Several studies have linked *5-HTT*LPR to psychopathology and psychotherapy outcomes, though with diverging conclusions (as reviewed in Schiele, Reif, et al., 2021). However, this meta-analysis concluded that, overall, the hypothesis of *5-HTT*LPR as a predictive marker for psychotherapy outcome in anxiety disorders lacks support. In light of this inconclusiveness, the present study aims to further investigate the role of *5-HTT*LPR in predicting treatment response, as understanding its potential influence could contribute to more personalized therapeutic approaches.

## 2 Experimental Procedures

### Sample

N=194 patients with spider phobia were drawn from a larger sample recruited within the framework of the transregional collaborative research center (CRC-TRR58) “Fear, Anxiety, Anxiety Disorders” funded by the German Research Foundation based on available *5-HTT*LPR/rs25531 genotype information (see table 1 for sample characteristics). The study was conducted at the University of Münster at the University Hospital of Würzburg, Germany. The study protocol was reviewed and approved by the respective ethics committees and corresponds to the ethical guidelines of the Declaration of Helsinki (1964). It has been registered at ClinicalTrials.gov with the identifier NCT03208400 and published by Schwarzmeier and Leehr et al. (2020). Written informed consent was obtained from all participants.

**Table 1.** Sample Characteristics.

|  | Total Sample | Genotype (5-HTTLPR/rs25531) |  | Comparison |  |
| --- | --- | --- | --- | --- | --- |
| | | High-expression | Low-expression | $\chi^2/t$-Test (df) | *p* |
| Sample size [n] | 194 | 71 | 123 |  |  |
| Female [*n* (%)] | 171(88.14%) | 62 (87.32%) | 108 (87.81%) | <0.01 (1) | 1 |
| Age (M±SD) | 28.25 (8.99) | 29.82 (8.88) | 27.44 (9.03) | 3.16 (192) | .076 |
| Age of Onset (M±SD) | 6.82 (4.51) | 6.74 (4.58) | 6.83 (4.50) | 0.02 (188) | .895 |
| BAT pre (M±SD) | 167.94 (66.71) | 159.25 (66.92) | 173.24 (66.65) | 1.98 (192) | .161 |
| SPQ pre (M±SD) | 22.88 (2.18) | 22.66 (2.16) | 23.00 (2.21) | 1.07 (192) | .302 |

*Note.* M= Mean; SD = standard deviation; BAT = Behavioral Avoidance Task; SPQ = Spider Phobia Questionnaire; high-expression genotype: L<sub>A</sub>/L<sub>A</sub>; low-expression genotypes: S/S, S/L<sub>G</sub>, L<sub>G</sub>/L<sub>G</sub>, S/L<sub>A</sub>, L<sub>G</sub>/L<sub>A</sub>.

The main inclusion criterion for the study was a diagnosis of spider phobia as assessed with the structured clinical interview for DSM-IV (SCID, Wittchen, 1997) and the Spider Phobia Questionnaire (SPQ, Klorman et al., 1974) with a cut-off score of 20 and above. Participants had to be between 18 and 65 years old, right-handed, fluent in German, and of Caucasian descent (self-report up to two generations). Spider phobia had to be the primary diagnosis; other specific phobias of the animal subtype as well as mild to moderate depression were allowed if less pronounced. Participants suffering from a lifetime diagnosis of other mental disorders as well as receiving concurrent (psycho-) pharmacological or psychotherapeutic treatment were excluded.

### Experimental Procedure

For an extensive description of study procedure and treatment protocol, see Schwarzmeier and Leehr et al. (2020). In short, patients underwent a one-session exposure treatment presented in VR. At baseline (pre-VRET), after VRET (post), and at a 6-month follow-up (FU), behavioral symptom severity was assessed with a standardized behavioral avoidance test (BAT), with a reduction of more than 50% defined a priori as response. Self-reported symptom severity was assessed with the SPQ, with a reduction of 30% defined a priori as response.

The standardized VRET was conducted by trained psychologists. A total of five scenarios from the program VTplus Research Systems (VTplus GmbH, Würzburg) were selected to confront the participants with different types of spiders in different situations. Before, during and after the scenarios, participants were asked to rate their fear on a scale from 0 (no fear) to 100 (maximum imaginable fear) at multiple time points. Each scenario defined anchor points which every participant was supposed to reach. To successfully complete the scenario, these anchor points had to be reached and the fear rating had to be either below 20 or stagnating for the third time point in a row. Maximum duration of the whole VRET appointment was 2.5 hours (mean= 81 minutes, sd= 25.69).

### Genotyping

Two 9 ml ethylenediamine tetraacetic acid (EDTA) monovettes of venous blood were drawn at baseline and immediately frozen at minus 80°C until further processing. Deoxyribonucleic acid (DNA) was then isolated using the FlexiGene DNA Kit (QIAGEN, Hilden, Germany). The genotyping for *5-HTT*LPR and the functionally related single nucleotide polymorphism rs25531 was conducted according to published protocols (Wendland et al., 2006). The Hardy-Weinberg criterion, as calculated by using the online program DeFinetti (Institute of Human Genetics, Helmholtz Zentrum München; ihg.gsf.de), was fulfilled (exact test; *p* = .732). Based on their genotype information, participants were grouped into high-(L_A_/L_A_), and low-expression (S/S, S/L_G,_L_G_/L_G_, S/L_A,_ L_G_/L_A_).

### Statistical Analyses

Statistical analyses were performed using the software *R*. To investigate the effect of *5-HTT* genotype on treatment response, chi-square tests were performed, with a significance level of *p* < .05.

## 3 Results

Clinical results are reported elsewhere (Leehr et al., 2021). In short, VRET significantly decreased symptom severity on both outcome measures. Table 2 displays the frequency distributions for behavioral responders and non-responders according to genotype in the present sample. There was a significant association between genotype and behavioral response after treatment (*χ*^2^ (1) =9.99, *p* = .002, Effect Size Cramer’s *V* = 0.227). Participants with the high-expression genotype showed higher rates of treatment response in comparison to those with low-expression genotypes. At the 6-month follow-up, the association between genotype and treatment response remained significant (*χ*^2^ (1) =5.06, *p* = .024, Effect Size Cramer’s *V* = 0.162). While the non-responder-to-responder ratio decreased over time for both groups, indicating improved treatment outcome over time, high-expression genotype carriers continued to have better response outcome compared to low-expression genotype carriers (see Figure 1 for a visual depiction).

**Table 2.** Cross Table for Behavioral Response by Genotype.

|  | Frequencies Post [FU] |  | Ratio Post [FU] |  |
| --- | --- | --- | --- | --- |
|  | NR | R | NR:R | Total |
| **High-expression** | 26 [22] | 45 [49] | 0.58 [0.45] | 71 [71] |
| **Low-expression** | 74 [58] | 49 [64] | 1.51 [0.91] | 123 [122] |
| Total | 100 [80] | 94 [113] | 1.06 [0.71] | 194 [193] |

*Note.* Frequencies of responders (R) and non-responders (NR) according to behavioral symptom reduction post-treatment [and at the 6 months follow-up, FU] grouped by high-expression (L<sub>A</sub>L<sub>A</sub>) and low-expression genotype (S/L<sub>A</sub>, L<sub>G</sub>/L<sub>A</sub> S/S, S/L<sub>G</sub>, L<sub>G</sub>/L<sub>G</sub>).

**Figure 1.**
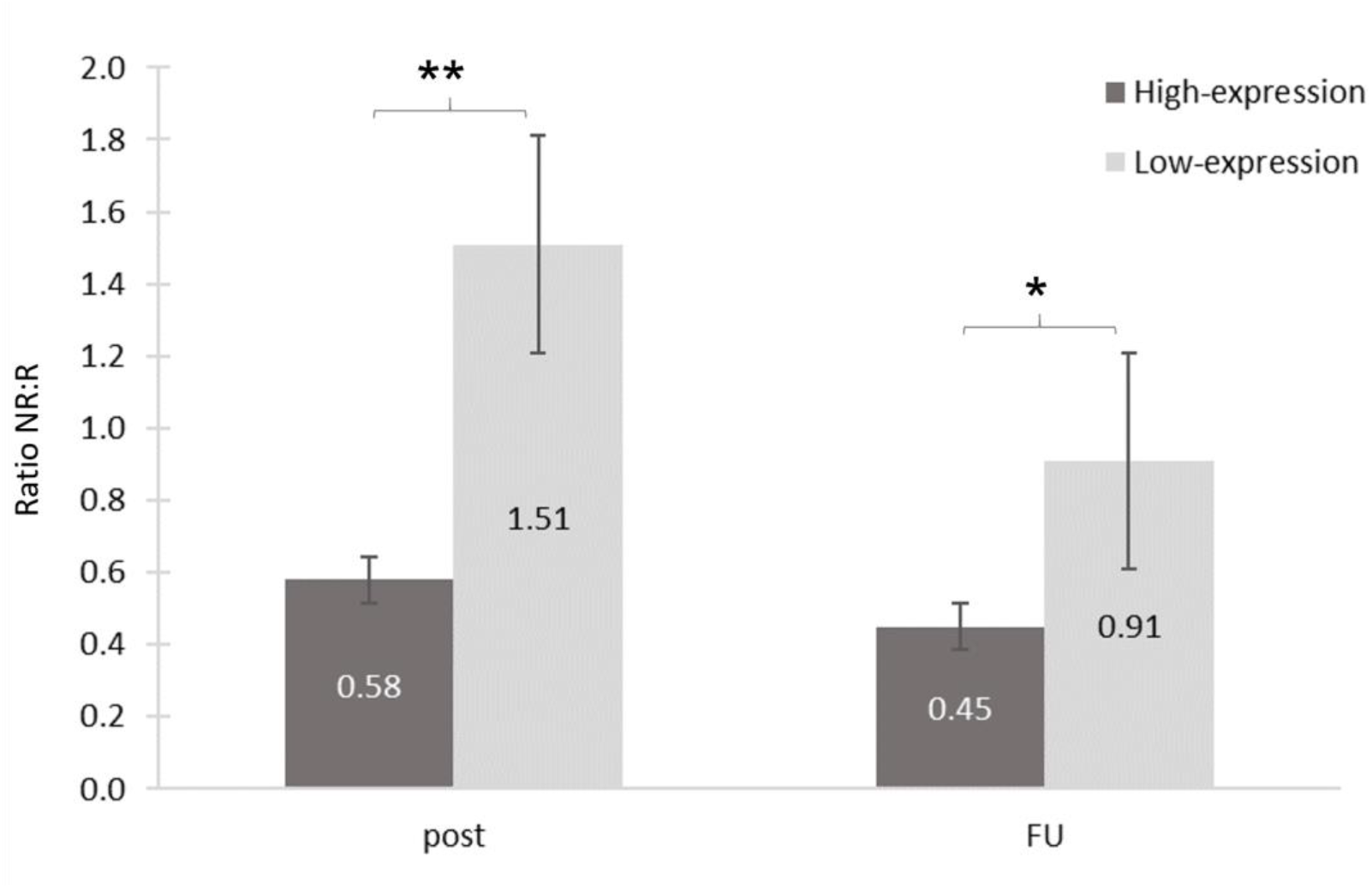
Ratio of non-responders (NR) to responders (R) (NR:R) according to BAT for high-expression (L_A_L_A_) and low-expression genotype (S/L_A,_ L_G_/L_A_ S/S, S/L_G,_ L_G_/L_G_) at the two time points: post-treatment, and 6-month follow-up (FU). \*\**p* < .01 \**p* < .05. Error bars depict standard error.

No association was found between genotype and self-reported symptom severity measured with the SPQ (short term: *χ*^*2*^ (1) = 0.59, *p* = .442, long term: *χ*^*2*^ (1) = 1.57, *p* =.210).

## 4 Discussion

In the present study, *5-HTT*LPR/rs25531 high-expression (L_A_L_A_) genotype carriers demonstrated better behavioral treatment outcome compared to low-expression genotype groups. With a non-responders-to-responders ratio of 0.58, L_A_L_A_ carriers were almost twice as likely to respond to the exposure treatment as to not respond. Conversely, low-expression genotypes carriers were 1.5 times as likely to show non-response as to show response. Although outcome rates improved over time for both groups, the high-expression genotype was still associated with superior long-term treatment outcome.

These findings mirror the heterogeneity of previous research and stand in opposition to a meta-analysis reporting no significant association between *5-HTT*LPR and CBT outcomes in anxiety disorders (Schiele, Reif, et al., 2021). Apart from potential differences in therapygenetic effects across anxiety disorders, discrepancies might be explained by the differential susceptibility hypothesis. This framework suggests that genetic variants may not directly confer vulnerability to negative environmental influences, but instead increase susceptibility to environmental influences of both positive and negative nature (Belsky et al., 2009). The *5-HTT*LPR/rs25531 L_A_L_A_ genotype, in particular, has been proposed to exert an either unfavorable or protective effect on anxiety depending on the quality of environmental input (Schiele et al., 2016). Accordingly, L_A_L_A_ genotype carriers may respond more favorably to CBT due to higher susceptibility to positive environmental influence, including therapeutic interventions. Indeed, carriers of the *5-HTT*LPR L-allele appear to exhibit facilitated fear extinction, a key mechanism in exposure-based therapies (for meta-analysis, see Miño et al., 2023). Moreover, these results are consistent with studies linking the high expression *5-HTT*LPR LL or *5-HTT*LPR/rs25531 L_A_L_A_ genotypes to better response to pharmacological treatment (Porcelli et al., 2012). On the neurobiological level, *5-HTT*LPR genotype seems to modulate amygdala activation in response to negative cues, hinting at a plausible mechanism through which genetic variation shapes susceptibility to environmental input and ultimately influences treatment response (Dannlowski et al., 2010).

Interestingly, in contrast to differences in behavioral treatment response, we did not find a genotype effect on self-reported fear ratings as measured with the SPQ. This might be due to more complex and harder-to-isolate interactions between genes and the environment when it comes to cognition (Tucker-Drob, Briley & Harden, 2013). While behavioral measures capture observable fear responses, self-reports rely on introspection and are shaped by different factors such as cognitive biases and social influences. This may obscure genetic effects, making them less detectable in subjective assessments compared to more automatic behavioral responses, such as avoidance behavior. Since extinction learning is a key mechanism in the treatment of anxiety disorders, the behavioral avoidance test may be more suitable than a self-report measure to capture this learning outcome. Accordingly, differences in how treatment response is measured across studies may be a further contributor to the heterogeneity of therapygenetic findings in the literature.

There are several limitations that need to be considered when interpreting our results. First of all, the generalizability of our findings might be limited by the rather specific context of our study. As outlined above, the moderate genotype effect (*V*=0.23) found in our homogenous sample of spider phobic participants in response to a highly standardized VRET might not hold up under more externally valid circumstances including more confounding factors (Schiele, Reif, et al., 2021).

Furthermore, the candidate gene approach used in our study has received growing criticism in past years, particularly in light of smaller sample sizes. By focusing on hypothesis-driven, select polymorphisms, the candidate gene approach offers limited genetic coverage, missing the broader polygenic influences and neglecting considerations of gene-environment interactions and epigenetics (Assary et al., 2018). The criticism has prompted a shift toward more comprehensive and unbiased methods like genome-wide association studies (GWAS), which might be better suited to capture the complex, multifactorial nature of mental disorders. Despite these limitations, the candidate gene approach might be advantageous for its focused hypothesis and mechanism testing, allowing researchers to investigate specific genes with the potential to directly identify significant genetic contributions, potentially leading to targeted interventions or treatments.

Finally, epigenetic mechanisms such as DNA methylation or histone modifications (Schiele et al., 2020) play a crucial role in regulating gene function and have been associated with therapy outcomes. For instance, *5-HTT* methylation has been shown to predict response to both antidepressant treatment (Schiele, Zwanzger, et al., 2021) as well as exposure therapy (Schiele, Thiel, et al., 2021).

To summarize, the present findings support the assumption of genotype-dependent treatment outcome in spider phobia, with *5-HTT*LPR/rs25531 high-expression genotype carriers showing superior behavioral treatment outcome to a standardized exposure treatment than low-expression genotypes. Conclusions drawn for therapy could be to offer rather individualized treatment approaches to carriers of the low-expressing genotype in order to improve response. Further research should independently replicate this finding, expand it to different anxiety disorders and type of treatments, and should take into account interacting and modulating factors especially with regard to (epi)genetic and environmental influences.

### Declaration of generative AI and AI-assisted technologies in the writing process

During the preparation of this work the author(s) used DeepL and ChatGPT in order to improve language and readability. After using this tool/service, the author(s) reviewed and edited the content as needed and take(s) full responsibility for the content of the publication.

## Data Availability

Due to the sensitive nature of the data, which includes genetic information, individual-level data cannot be shared in order to protect participant privacy. However, aggregated data are available from the corresponding author upon reasonable request.

